# Reduced Oxygen Extraction Fraction in Deep Cerebral Veins Associated with Cognitive Impairment in Multiple Sclerosis

**DOI:** 10.1101/2024.01.10.24301049

**Authors:** Hasan Sawan, Chenyang Li, Sagar Buch, Evanthia Bernitsas, E. Mark Haacke, Yulin Ge, Yongsheng Chen

**Author notes:** **Corresponding author** Yongsheng Chen, PhD., Address: 4201 St. Antoine, UHC-8D, Detroit, MI 48098, USA.

## Abstract

Studying the relationship between cerebral oxygen utilization and cognitive impairment is essential to understanding neuronal functional changes in the disease progression of multiple sclerosis (MS). This study explores the potential of using venous susceptibility in internal cerebral veins (ICVs) as an imaging biomarker for cognitive impairment in relapsing-remitting MS (RRMS) patients. Quantitative susceptibility mapping derived from fully flow-compensated MRI phase data was employed to directly measure venous blood oxygen saturation levels (S_v_O_2_) in the ICVs. Results revealed a significant reduction in the susceptibility of ICVs (212.4 ± 30.8 ppb vs 239.4 ± 25.9 ppb) and a significant increase of S_v_O_2_ (74.5 ± 1.89 % vs 72.4 ± 2.23 %) in patients with RRMS compared with age- and sex-matched healthy controls. Both the susceptibility of ICVs (*r* = 0.646, *p* = 0.004) and the S_v_O_2_ (*r* = −0.603, *p* = 0.008) exhibited a strong correlation with cognitive decline in these patients assessed by the Paced Auditory Serial Addition Test, while no significant correlation was observed with clinical disability measured by the Expanded Disability Status Scale. The findings suggest that venous susceptibility in ICVs has the potential to serve as a specific indicator of oxygen metabolism and cognitive function in RRMS.

## INTRODUCTION

Multiple sclerosis (MS) is an inflammatory and neurodegenerative disease characterized by neural tissue atrophy and multifocal inflammatory and demyelinating lesions in the central nervous system.^1^ The cause and course of MS lesions are not fully elucidated, with neurodegeneration emerging as the primary driving force behind both disease progression and cognitive decline.^2^ Several hypotheses underlie progressive neurodegeneration in MS. One explanation revolves around energy deficiency in lesions, termed virtual (also known as metabolic or histotoxic) hypoxia.^3^ In contrast to “true” hypoxia, virtual hypoxia does not necessarily involve reduced oxygen availability; rather, it is characterized by diminished oxygen utilization with mitochondrial dysfunction. Virtual hypoxia likely plays a significant role in the pathophysiology of neurodegeneration in MS.^4^ This effect is not just localized to lesions; reduced oxygen extraction fraction (OEF), a marker of cell degeneration or dysfunction, has been described in normal-appearing tissue, including in deep and cortical gray matter of MS patients.^5, 6^

Furthermore, a reduced cerebral metabolic rate of oxygen (CMRO_2_), a similar measure of OEF, has long been related to symptomatology of MS, particularly cognitive impairment.^6–9^ Cognitive impairment in MS can arise from decreased oxygen metabolism manifesting either as direct damage to brain tissue or as a decline in neuronal/axonal activity.^10–12^ Specifically, cognitive dysfunction is linked to damage in deep gray matter nuclei^11, 13, 14^, with the thalamus being identified as the primary contributor.^15, 16^ Therefore, measurements of OEF in deep cerebral regions could serve as a specific imaging biomarker of cognitive decline in MS.

Studies on cerebral oxygen metabolism have traditionally relied on positron emission tomography (PET), the gold-standard, for assessment of CMRO_2_ in MS.^7, 9, 17^ However, PET is disadvantaged clinically by exposure to radiation and relatively limited availability. Recently, non-invasive MRI-based methods have emerged as a major focus for the quantification of OEF and CMRO_2_.^18–20^ Various techniques measuring cerebral OEF have been proposed, including phase or quantitative susceptibility mapping (QSM) based methods^21–23^ and a T2-relaxation-under-spin-tagging (TRUST) method^24^ for measuring macrovascular OEF; respiratory challenge calibrated blood-oxygen-level-dependent (BOLD) methods^25–27^; calibration-free quantitative BOLD based methods^28–30^; QSM-based voxel-wise OEF mappings^31–33^; as well as methods combining quantitative BOLD and QSM^34–36^. *Ge et al.* observed a significantly decreased OEF in MS patients measured in the superior sagittal sinus (SSS) using the TRUST method.^8^ The reduction of OEF was correlated with disease severity and total lesion load.^8^ *Fan et al*. also reported a reduction of cortical OEF in MS, using a phase-based method measuring pial veins parallel to the main field.^6^ More recently, *Cho et al.* utilized a voxel-wise OEF method combining QSM and quantitative BOLD, observing a whole brain region reduced OEF in MS patients.^5^ Common among these various methods is the use of venous blood susceptibility as a direct measure of venous oxygen saturation levels (S_v_O_2_) and OEF of the veins draining the cerebral tissue. QSM also eliminates the orientation-based errors in phase-based methods, which can accurately measure susceptibility predominantly for veins parallel to the main field.^37^ To its advantage, the venous QSM-based OEF method is readily available in clinical settings with minimal requirement for image post-processing.^37, 38^ However, it remains largely unknown whether the susceptibility of cerebral veins can be used as an imaging biomarker of neuronal activity in MS. Deep cerebral veins such as internal cerebral veins (ICVs) drain the periventricular white matter (WM) where most MS lesions including the slowly expanding lesions occur^39^, as well as subcortical grey matter nuclei which are associated with cognitive function^11^.

The objective of this study was to investigate the susceptibility of ICVs and the subsequent S_v_O_2_ in patients with relapsing-remitting MS (RRMS), and their underlying correlations with a clinical measure of cognitive function and disease severity.

## METHODS

### Subjects and Clinical Assessments

Patients with RRMS and age-matched healthy controls (HC) were recruited from the Neurology clinic at Wayne State University and approved by the local institutional review board. Written consent was obtained from each participant. The inclusion and exclusion criteria were described in a previous publication using the quantitative MRI data acquired from the same cohorts.^40^ A neurologist performed clinical assessment for MS patients, including the Expanded Disability Status Scale (EDSS) for disease severity^41^ and the Paced Auditory Serial Addition Test (PASAT) for cognitive function.^42^

### Data Acquisition and Processing

The QSM data presented in this paper were acquired with an interleaved flow-rephased and flow-dephased gradient echo sequence^43^. The data used in this retrospective study was not published previously. The imaging parameters were as follows: TR = 20 ms, flip angle = 12°, two echoes of 12.5 ms acquired with interleaved TRs, and an acquisition resolution of 0.67 x 1.34 x 2.0 mm^3^. There were 80 axial slices covering the whole brain. The scan time was 4 min 53 sec. The interleaved sequence had a fully flow-compensated echo of 12.5 ms for a bright blood image, and a flow-dephased echo of 12.5 ms for a dark blood image (Fig. 1A and 1B). The subtraction of these two magnitude images yielded an MR angiogram and venogram (MRAV) with no background tissue remaining (Fig. 1C)^43^. The phase of the flow-compensated echo was used for the reconstruction of the QSM data as described (Fig. 1D)^43^. In short, a quality-guided 3D best path algorithm, 3DSRNCP^44^, was used for phase unwrapping followed by background field removal using the SHARP algorithm with a kernel size of 8 pixels and a deconvolution threshold of 0.05^45^. Then, a truncated k-space division (TKD) was used to solve the inverse problem and to generate an initial susceptibility map^21^. Finally, an iterative algorithm with 4 iterations and a threshold-based geometry mask indicating major veins and high-susceptibility deep grey matter nuclei was used to reduce the streaking artifacts on the TKD-generated QSM.^46^ The final iterated QSM data improves the accuracy of the susceptibility for major veins, such as the ICVs.^46^

**Figure 1.**
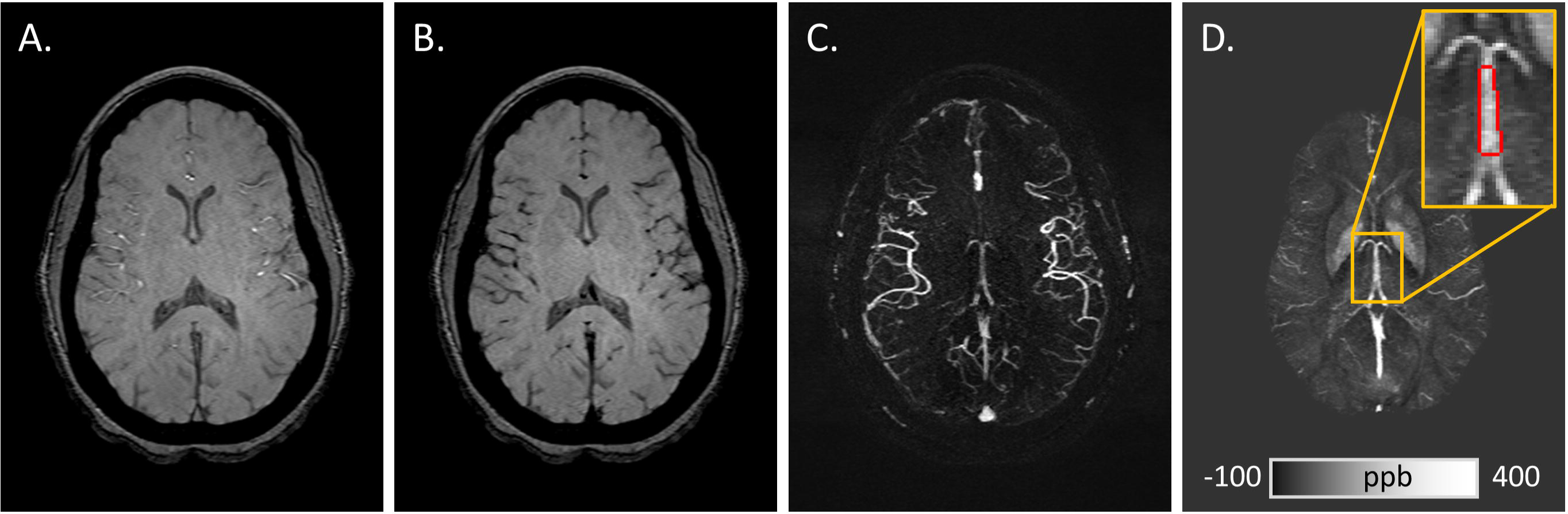
Representative images and regions-of-interest (ROI) for the internal cerebral veins (ICVs). The sequence provides a fully flow-compensated bright blood image (A) and a flow-dephased dark blood image (B). The subtraction of the two magnitude images gives MRAV data (C). The phase of the flow-compensated echo was used for reconstructing the QSM data (D). The ROI of the ICVs is shown as the red boundary in the inset (D).

### Data Analysis

Two observers independently traced the ICVs using a semi-automated region-growing tool in SPIN software (V1.5, MR Innovations Inc, Detroit, MI) on a maximum intensity projection (MIP) over 8 slices of the QSM data (Fig. 1D). To investigate the inter-rater and intra-rater reliability, each observer delineated the region-of-interest (ROI) of the ICVs again on all data a week after the initial delineation. To minimize the partial volume effect (PVE), the ROI was placed at the middle segment of the ICVs between the thalamostriate veins and medial atrial veins (Fig. 1D). Patient disease status was blinded to the observers during ROI delineation. The median susceptibility value of all pixels within the ROI (Δχ_ICV_) was used for each subject. Subsequently, the χ_ICV_ was converted to S_v_O_2_ using Eq 1.

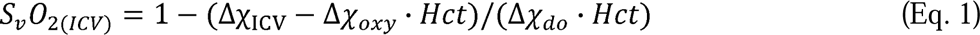

where Δχ_ICV_ is the measured susceptibility of the ICVs relative to water, Δχ_oxy_ is the susceptibility difference between fully oxygenated blood and water (−0.03 × 4π ppm), Δχ_do_ is the susceptibility difference between fully oxygenated and deoxygenated blood (0.27 × 4π ppm), and *Hct* is the average hematocrit expected for women (0.43) and men (0.45).^47, 48^

Statistical analysis was performed using MATLAB (R2017b, MathWorks, Natick, MA) with in-house developed scripts. The Wilcoxon rank sum test was used to compare the age, gender percentage, Δχ_ICV_, and S_v_O_2_ between the two cohorts. Inter-rater reliability of Δχ_ICV_ was assessed by the intraclass correlation coefficient (ICC) analysis^49^ and linear regression. Linear regression was used to correlate Δχ_ICV_with PASAT and EDSS scores in MS patients. *P* < 0.05 was considered statistically significant.

## RESULTS

The study enrolled 20 patients with RRMS and 10 age-matched HCs. Two MS patients’ data were excluded from the present study due to strong susceptibility artifacts caused by calcification in the pineal gland (around which the ICVs bifurcate from the great cerebral vein). The demographics and clinical assessments are listed in Table 1. The age and gender percentage were comparable between MS and HC. The reliability analysis demonstrated an excellent intra-rater (ICC = 0.97, *r*^2^ = 0.97, *p* < 0.001) and inter-rater (ICC = 0.96, *r*^2^ = 0.97, *p* < 0.001) agreement of the measuring method. Compared with HCs, patients with MS had a significantly reduced Δχ_ICV_(212.4 ± 30.8 ppb in MS vs 239.4 ± 25.9 ppb in HC), and a significantly increased S_v_O_2_ (74.5 ± 1.89 % in MS vs 72.4 ± 2.23 % in HC), as shown in Fig. 2A and 2B. The PASAT score, a measure of cognitive function, was strongly correlated with the Δχ_ICV_(*r* = 0.646, *p* = 0.004) and the S_v_O_2_ (*r* = −0.603, *p* = 0.008) in MS patients (Fig. 2C and 2D). In contrast, the disease severity score, EDSS, had no correlation effect with Δχ_ICV_ (*r* = −0.295, *p* = 0.236), nor S_v_O_2_ (*r* = 0.279, *p* = 0.260) in MS patients.

**Figure 2.**
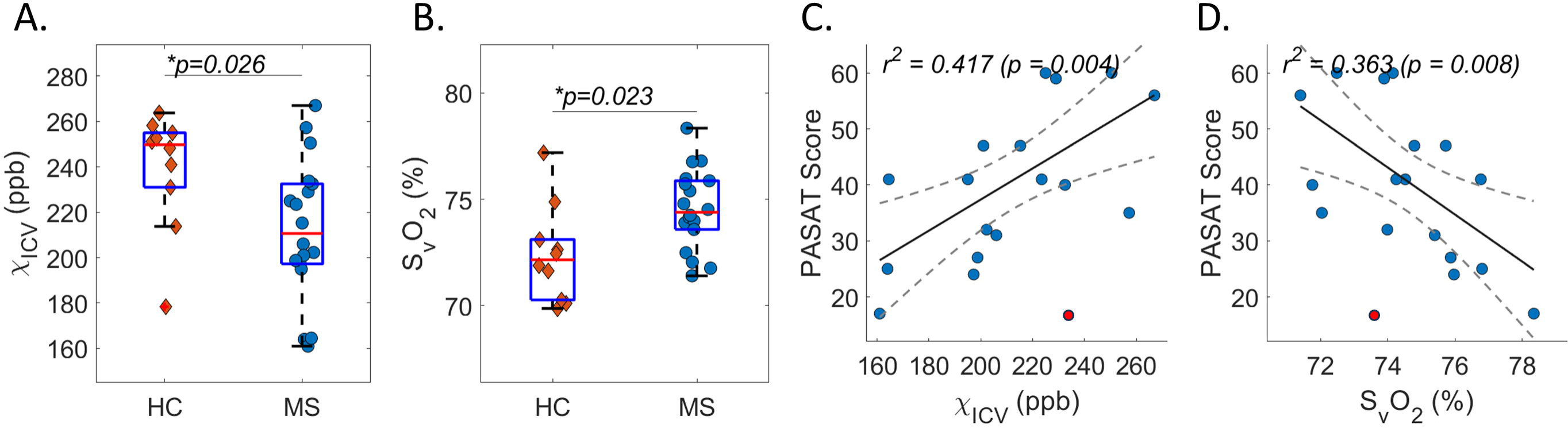
Increased deep cerebral venous blood oxygenation was correlated with cognitive decline. The box plot analysis shows a reduced Δχ_ICV_ (A) and increased S_v_O_2_ (C) in MS compared with healthy controls (HC). The PASAT score was correlated with Δχ_ICV_ (C) and S_v_O_2_ (D) in MS patients. One outliner with the lowest PASAT score (red data points on C and D) was excluded during the correlation analyses.

## DISCUSSION

In this study, we observed a reduced susceptibility of ICVs in RRMS patients compared to age- and sex-matched controls, indicating decreased deoxyhemoglobin levels in venous blood in ICVs. Consequently, there was an increased S_v_O_2_ in these patients, implicating a decreased OEF in the corresponding cerebral regions draining into the ICVs. These imaging biomarkers were correlated with cognitive impairment in these MS patients, as measured by PASAT. The high intra- and inter-rater ICC scores affirm the repeatability and reproducibility of the method.

Oxygen molecules bind to hemoglobin by eliminating unpaired electrons, forming diamagnetic oxyhemoglobin. Upon the release of oxygen from oxyhemoglobin in neuronal tissue, deoxyhemoglobin in venous blood becomes paramagnetic, resulting in relatively strong magnetic field perturbations in MRI. Therefore, the susceptibility of venous blood can serve as a direct measure of S_v_O_2_ given the negligible effect of other sources of susceptibility changes in the blood^48^. The phase accumulation as measured for a given echo time in a gradient echo sequence is proportional to the susceptibility changes. Indeed, phase images have been used to directly measure the S_v_O_2_ of pial veins parallel to the main field, where there is no magnetic dipole inversion.^6, 21, 22^ In this regard, QSM reconstructed from phase data makes it possible to measure venous blood oxygenation in all orientations^21, 37, 38^. However, the typically used multi-echo fitting based QSM often acquires the gradient echo data with a very short first TE and small echo spacing, making it impractical to use flow compensation in all gradient directions.^37, 50^ Uncompensated blood flow thereby becomes the major source of error in venous-QSM based OEF measurement.^23^ In addition, gradient echo sequences in clinical MRI scanners only have flow compensation options for the first echo, but not for the second and subsequent echoes. To overcome these issues, we used a single-echo based QSM method along with flow compensation in all gradient directions to minimize blood flow induced phase errors.^43, 46^ Furthermore, we used an optimal echo time of 12.5 ms, which is a fat-water in-phase echo time close to T_2_*/2 of the venous blood, to give sufficient signal-to-noise ratio (SNR) of the venous blood phase while also avoiding intravoxel aliasing of the data coming from the edges of veins.^37^ Although the present study used the QSM data acquired from a customized interleaved MRAV sequence, an identical equivalent can be implemented on clinical scanners using the product gradient echo sequence with a scan time of 2 min 27 sec by eliminating the interleaved TR for the subtraction-based MRAV.^43^

The observed increased S_v_O_2_ in the ICVs is likely due to reduced oxygen consumption in the deep cerebral regions that drain into the ICV, which include periventricular WM and deep gray matter regions. The superior thalamostriate veins, which drain the corpus striatum and thalamus, unite with the superior choroid veins, which drain the hippocampus, fornix, and corpus callosum, to become the ICVs.^51^ Damage to any of these deep gray matter nuclei are implicated in cognitive impairment in MS.^11, 13–16^ The association found in this study between increased S_v_O_2_ and cognitive impairment suggests that reduced oxygen consumption, possibly due to virtual hypoxia, is a factor in the damage to these regions. This further aligns with previous studies that found reduced CMRO_2_ in deep gray matter nuclei.^5, 6^ In addition, reduced oxygen consumption in deep cerebral gray matter may play a key role in cognitive function in other neurodegenerative disorders such as Alzheimer’s disease.^52^ WM damage is also of note, as the subependymal veins, which drain the periventricular WM regions through medullary veins, in turn drain into the ICVs. The periventricular region where MS lesions are most commonly seen may also play a role in explaining S_v_O_2_ increases in the ICVs. Previous studies have explored the role of WM and reported a contribution from WM to MS-related cognitive impairment.^53, 54^ This is also in line with a previous study demonstrating diminished medullary vein visibility in periventricular WM territories in MS patients.^55^ Taken together, the findings of this study emphasize the utility of deep cerebral venous susceptibility as a potential imaging biomarker of cognitive decline in RRMS as well as normal aging and other neurodegenerative disorders.

There are several limitations to this study. First, this retrospective study includes only a small cohort of RRMS patients. While RRMS patients constitute most MS patients^56^, limiting the cohort to a single MS subtype may miss an opportunity to explore varying profiles of pathology, given that the prevalence of cognitive impairment varies widely among subtypes.^57^ Second, while χ_ICV_ and S_v_O_2_ are good measures of venous deoxyhemoglobin and oxygenation, they fall short of being a complete stand-in for OEF as understood by Fick’s principle, which also requires a measurement of arterial blood oxygen saturation supplying the same tissue. Although arterial oxygen saturation levels can be measured by a finger pulse oximeter^8^, this data was not collected from the present cohort when they underwent MRI. Importantly, true hypoxia plays an equally prevalent role in MS^4^, and previous studies have found lowered deep gray matter blood oxygen saturation in MS using infrared spectroscopy — reasonably, oxygen saturation in the finger may vary from that in the cerebrum.^58^ Further studies would benefit from measuring arterial blood oxygen saturation, ideally in the counterpart arteries to the ICVs. Third, QSM reconstruction has a step of brain extraction which typically discards cerebral edge regions including most of the cortical veins and the SSS.^37^ Thus, the current method is limited to measuring only from veins available in the QSM data. This could be circumvented in future studies by using whole brain QSM methods.^59, 60^ Fourth, PASAT alone may not be the most reliable indicator of our participants’ cognitive function. Despite its high test-retest reliability and internal consistency, it is influenced by patients’ mathematical abilities, intelligence quotient, and age.^42^ Finally, significant PVE of the ICVs was observed by the generally lower venous susceptibility compared to the theoretical value of 450 ppb.^37^ The PVE could be reduced by using a higher imaging resolution. Practically speaking, acquiring a single echo with full flow compensation and a resolution of 0.5 × 0.5 × 1.5 mm³ could be achieved in less than 5 minutes on clinical MRI scanners. This could be accomplished by using a TE of 12.5 ms, a TR of 20 ms, a low bandwidth of 150 Hz/pixel to ensure the SNR of the data, a parallel imaging acceleration factor of 2, and 80 axial slices covering most of the brain. Further studies should consider the correction of PVE in QSM reconstruction. Nevertheless, the findings of this study are endorsed by the excellent agreement of the inter-rater and intra-rater analyses, and the factor that the exact same method was used on patients and controls.

In summary, this study demonstrates the feasibility of using deep cerebral venous susceptibility as an imaging biomarker of cognitive impairment in MS. This pilot study warrants further investigation of cerebral venous QSM as a clinically feasible tool in assessing cerebral oxygen metabolism in large-scale prospective clinical studies in MS and other neurodegenerative disorders.

**Table 1.** Demographics and data. Data were presented as mean ± standard deviation (minimum–maximum)

|  | MS Patients (n = 18) | Controls (n = 10) | *p*-value |
| --- | --- | --- | --- |
| Age (years) | 36.7 ± 7.7 (24 – 54) | 35.6 ± 12.7 (21 – 58) | 0.262 |
| Male Percentage (%) | 27.8 | 40.0 | 0.534 |
| $\chi_{ICV}$ (ppb) | 212.4 ± 30.8 (161.0 – 267.0) | 239.4 ± 25.9 (178.5 – 263.8) | 0.026* |
| $S_vO_2$ (%) | 74.5 ± 1.89 (71.4 – 78.3) | 72.4 ± 2.23 (69.9 – 77.2) | 0.023* |
| PASAT | 38.8 ± 14.3 (15.0 – 60.0) | - | - |
| EDSS | 5.7 ± 4.2 (1.0 – 17.0) | - | - |

## Data Availability

All data produced in the present study are available upon reasonable request to the corresponding author.

## Acknowledgment

We thank Fahad Malik for his contribution to the inter-rater reliability analysis. Research reported in this publication was partially supported by the NIH/NINDS under award number R61NS119434. The content is solely the responsibility of the authors and does not necessarily represent the official views of the NIH. This work was also partially supported by the Office of the Vice President for Research at Wayne State University through their support of the MR Research Facility. The research data collection was supported by AbbVie Inc through grant award number C91205.

## REFERENCES

1. Grigoriadis N and Van Pesch V. A basic overview of multiple sclerosis immunopathology. European journal of neurology. 2015; 22: 3–13.

2. Lucchinetti C, Brück W, Parisi J, Scheithauer B, Rodriguez M and Lassmann H. Heterogeneity of multiple sclerosis lesions: implications for the pathogenesis of demyelination. Annals of Neurology: Official Journal of the American Neurological Association and the Child Neurology Society. 2000; 47: 707–17.

3. Trapp BD and Stys PK. Virtual hypoxia and chronic necrosis of demyelinated axons in multiple sclerosis. The Lancet Neurology. 2009; 8: 280–91.

4. Halder SK and Milner R. Hypoxia in multiple sclerosis; is it the chicken or the egg? Brain. 2021; 144: 402–10.

5. Cho J, Nguyen TD, Huang W, et al. Brain oxygen extraction fraction mapping in patients with multiple sclerosis. Journal of Cerebral Blood Flow & Metabolism. 2022; 42: 338–48.

6. Fan AP, Govindarajan ST, Kinkel RP, et al. Quantitative oxygen extraction fraction from 7-Tesla MRI phase: reproducibility and application in multiple sclerosis. Journal of Cerebral Blood Flow & Metabolism. 2015; 35: 131–9.

7. Brooks D, Leenders K, Head G, Marshall J, Legg N and Jones T. Studies on regional cerebral oxygen utilisation and cognitive function in multiple sclerosis. Journal of neurology, neurosurgery, and psychiatry. 1984; 47: 1182.

8. Ge Y, Zhang Z, Lu H, et al. Characterizing brain oxygen metabolism in patients with multiple sclerosis with T2-relaxation-under-spin-tagging MRI. Journal of Cerebral Blood Flow & Metabolism. 2012; 32: 403–12.

9. Sun X, Tanaka M, Kondo S, Okamoto K and Hirai S. Clinical significance of reduced cerebral metabolism in multiple sclerosis: a combined PET and MRI study. Annals of nuclear medicine. 1998; 12: 89–94.

10. Margoni M, Preziosa P, Rocca MA and Filippi M. Depressive symptoms, anxiety and cognitive impairment: emerging evidence in multiple sclerosis. Translational Psychiatry. 2023; 13: 264.

11. Rocca MA, Amato MP, De Stefano N, et al. Clinical and imaging assessment of cognitive dysfunction in multiple sclerosis. The Lancet Neurology. 2015; 14: 302–17.

12. Tozlu C, Olafson E, Jamison KW, et al. The sequence of regional structural disconnectivity due to multiple sclerosis lesions. Brain Communications. 2023; 5: fcad332.

13. Batista S, Zivadinov R, Hoogs M, et al. Basal ganglia, thalamus and neocortical atrophy predicting slowed cognitive processing in multiple sclerosis. Journal of neurology. 2012; 259: 139–46.

14. Muhlert N, Atzori M, De Vita E, et al. Memory in multiple sclerosis is linked to glutamate concentration in grey matter regions. *Journal of Neurology*, Neurosurgery & Psychiatry. 2014.

15. Houtchens M, Benedict R, Killiany R, et al. Thalamic atrophy and cognition in multiple sclerosis. Neurology. 2007; 69: 1213–23.

16. Minagar A, Barnett MH, Benedict RH, et al. The thalamus and multiple sclerosis: modern views on pathologic, imaging, and clinical aspects. Neurology. 2013; 80: 210–9.

17. Brier MR and Taha F. Measuring Pathology in Patients with Multiple Sclerosis Using Positron Emission Tomography. Current neurology and neuroscience reports. 2023; 23: 479–88.

18. Biondetti E, Cho J and Lee H. Cerebral oxygen metabolism from MRI susceptibility. NeuroImage. 2023; 276: 120189.

19. Jiang D and Lu H. Cerebral oxygen extraction fraction MRI: Techniques and applications. Magnetic resonance in medicine. 2022; 88: 575–600.

20. Rodgers ZB, Detre JA and Wehrli FW. MRI-based methods for quantification of the cerebral metabolic rate of oxygen. Journal of Cerebral Blood Flow & Metabolism. 2016; 36: 1165–85.

21. Haacke E, Tang J, Neelavalli J and Cheng Y. Susceptibility mapping as a means to visualize veins and quantify oxygen saturation. Journal of Magnetic Resonance Imaging. 2010; 32: 663–76.

22. Fan AP, Bilgic B, Gagnon L, et al. Quantitative oxygenation venography from MRI phase. Magnetic resonance in medicine. 2014; 72: 149–59.

23. Xu B, Liu T, Spincemaille P, Prince M and Wang Y. Flow compensated quantitative susceptibility mapping for venous oxygenation imaging. Magnetic resonance in medicine. 2014; 72: 438–45.

24. Lu H and Ge Y. Quantitative evaluation of oxygenation in venous vessels using T2_-_ relaxation_-_under_-_spin_-_tagging MRI. Magnetic Resonance in Medicine: An Official Journal of the International Society for Magnetic Resonance in Medicine. 2008; 60: 357–63.

25. Bulte DP, Kelly M, Germuska M, et al. Quantitative measurement of cerebral physiology using respiratory-calibrated MRI. Neuroimage. 2012; 60: 582–91.

26. Gauthier CJ and Hoge RD. A generalized procedure for calibrated MRI incorporating hyperoxia and hypercapnia. Human brain mapping. 2013; 34: 1053–69.

27. Wise RG, Harris AD, Stone AJ and Murphy K. Measurement of OEF and absolute CMRO2: MRI-based methods using interleaved and combined hypercapnia and hyperoxia. Neuroimage. 2013; 83: 135–47.

28. Yablonskiy DA and Haacke EM. Theory of NMR signal behavior in magnetically inhomogeneous tissues: the static dephasing regime. Magnetic resonance in medicine. 1994; 32: 749–63.

29. He X and Yablonskiy DA. Quantitative BOLD: mapping of human cerebral deoxygenated blood volume and oxygen extraction fraction: default state. Magnetic Resonance in Medicine: An Official Journal of the International Society for Magnetic Resonance in Medicine. 2007; 57: 115–26.

30. An H and Lin W. Quantitative measurements of cerebral blood oxygen saturation using magnetic resonance imaging. Journal of Cerebral Blood Flow & Metabolism. 2000; 20: 1225–36.

31. Zhang J, Cho J, Zhou D, et al. Quantitative susceptibility mapping_-_based cerebral metabolic rate of oxygen mapping with minimum local variance. Magnetic Resonance in Medicine. 2018; 79: 172–9.

32. Zhang J, Liu T, Gupta A, Spincemaille P, Nguyen TD and Wang Y. Quantitative mapping of cerebral metabolic rate of oxygen (CMRO2) using quantitative susceptibility mapping (QSM). Magnetic resonance in medicine. 2015; 74: 945–52.

33. Zhang J, Zhou D, Nguyen TD, Spincemaille P, Gupta A and Wang Y. Cerebral metabolic rate of oxygen (CMRO2) mapping with hyperventilation challenge using quantitative susceptibility mapping (QSM). Magnetic resonance in medicine. 2017; 77: 1762–73.

34. Cho J, Kee Y, Spincemaille P, et al. Cerebral metabolic rate of oxygen (CMRO2) mapping by combining quantitative susceptibility mapping (QSM) and quantitative blood oxygenation level_-_dependent imaging (qBOLD). Magnetic resonance in medicine. 2018; 80: 1595–604.

35. Cho J, Lee J, An H, Goyal MS, Su Y and Wang Y. Cerebral oxygen extraction fraction (OEF): Comparison of challenge-free gradient echo QSM+ qBOLD (QQ) with 15O PET in healthy adults. Journal of Cerebral Blood Flow & Metabolism. 2021; 41: 1658–68.

36. Cho J, Ma Y, Spincemaille P, Pike GB and Wang Y. Cerebral oxygen extraction fraction: comparison of dual_-_gas challenge calibrated BOLD with CBF and challenge_-_free gradient echo QSM+ qBOLD. Magnetic resonance in medicine. 2021; 85: 953–61.

37. Haacke EM, Liu S, Buch S, Zheng W, Wu D and Ye Y. Quantitative susceptibility mapping: current status and future directions. Magnetic resonance imaging. 2015; 33: 1–25.

38. Buch S, Ye Y and Haacke EM. Quantifying the changes in oxygen extraction fraction and cerebral activity caused by caffeine and acetazolamide. Journal of Cerebral Blood Flow & Metabolism. 2017; 37: 825–36.

39. Buch S, Subramanian K, Chen T, et al. Characterization of white matter lesions in multiple sclerosis using proton density and T1-relaxation measures. Magnetic Resonance Imaging. 2023.

40. Haacke EM, Bernitsas E, Subramanian K, et al. A Comparison of Magnetic Resonance Imaging Methods to Assess Multiple Sclerosis Lesions: Implications for Patient Characterization and Clinical Trial Design. Diagnostics. 2021; 12: 77.

41. Meyer-Moock S, Feng Y-S, Maeurer M, Dippel F-W and Kohlmann T. Systematic literature review and validity evaluation of the Expanded Disability Status Scale (EDSS) and the Multiple Sclerosis Functional Composite (MSFC) in patients with multiple sclerosis. BMC neurology. 2014; 14: 1–10.

42. Tombaugh TN. A comprehensive review of the paced auditory serial addition test (PASAT). Archives of clinical neuropsychology. 2006; 21: 53–76.

43. Chen Y, Liu S, Buch S, Hu J, Kang Y and Haacke EM. An interleaved sequence for simultaneous magnetic resonance angiography (MRA), susceptibility weighted imaging (SWI) and quantitative susceptibility mapping (QSM). Magnetic resonance imaging. 2018; 47: 1–6.

44. Abdul-Rahman HS, Gdeisat MA, Burton DR, Lalor MJ, Lilley F and Moore CJ. Fast and robust three-dimensional best path phase unwrapping algorithm. Applied optics. 2007; 46: 6623–35.

45. Schweser F, Deistung A, Lehr BW and Reichenbach JR. Quantitative imaging of intrinsic magnetic tissue properties using MRI signal phase: an approach to in vivo brain iron metabolism? Neuroimage. 2011; 54: 2789–807.

46. Tang J, Liu S, Neelavalli J, Cheng YC, Buch S and Haacke EM. Improving susceptibility mapping using a threshold-based K-space/image domain iterative reconstruction approach. Magn Reson Med. 2013; 69: 1396–407.

47. Spees WM, Yablonskiy DA, Oswood MC and Ackerman JJ. Water proton MR properties of human blood at 1.5 Tesla: Magnetic susceptibility, T1, T2, T, and non_-_Lorentzian signal behavior. Magnetic Resonance in Medicine: An Official Journal of the International Society for Magnetic Resonance in Medicine. 2001; 45: 533–42.

48. Weisskoff RM and Kiihne S. MRI susceptometry: image_-_based measurement of absolute susceptibility of MR contrast agents and human blood. Magnetic resonance in medicine. 1992; 24: 375–83.

49. Shrout PE and Fleiss JL. Intraclass correlations: uses in assessing rater reliability. Psychological bulletin. 1979; 86: 420.

50. Wang Y and Liu T. Quantitative susceptibility mapping (QSM): decoding MRI data for a tissue magnetic biomarker. Magnetic resonance in medicine. 2015; 73: 82–101.

51. Wang J, Wang J, Sun J and Gong X. Evaluation of the anatomy and variants of internal cerebral veins with phase-sensitive MR imaging. Surgical and radiologic anatomy. 2010; 32: 669–74.

52. Yang A, Zhuang H, Du L, et al. Evaluation of whole-brain oxygen metabolism in Alzheimer’s disease using QSM and quantitative BOLD. NeuroImage. 2023; 282: 120381.

53. Elkhooly M, Bao F, Raghib M, Millis S and Bernitsas E. Role of white matter in cognitive impairment among relapsing remitting multiple sclerosis patients. Multiple Sclerosis and Related Disorders. 2023; 79: 105030.

54. Engl C, Tiemann L, Grahl S, et al. Cognitive impairment in early MS: contribution of white matter lesions, deep grey matter atrophy, and cortical atrophy. Journal of Neurology. 2020; 267: 2307–18.

55. Ge Y, Zohrabian VM, Osa EO, et al. Diminished visibility of cerebral venous vasculature in multiple sclerosis by susceptibility_-_ weighted imaging at 3.0 Tesla. Journal of Magnetic Resonance Imaging: An Official Journal of the International Society for Magnetic Resonance in Medicine. 2009; 29: 1190–4.

56. Klineova S and Lublin FD. Clinical course of multiple sclerosis. Cold Spring Harbor perspectives in medicine. 2018; 8: a028928.

57. Ruano L, Portaccio E, Goretti B, et al. Age and disability drive cognitive impairment in multiple sclerosis across disease subtypes. Multiple Sclerosis Journal. 2017; 23: 1258–67.

58. Hubbard NA, Sanchez Araujo Y, Caballero C, et al. Evaluation of visual-evoked cerebral metabolic rate of oxygen as a diagnostic marker in multiple sclerosis. Brain Sciences. 2017; 7: 64.

59. Buch S, Chen Y and Haacke EM. Susceptibility mapping of the dural sinuses and other superficial veins in the brain. Magnetic resonance imaging. 2019; 57: 19–27.

60. Wei H, Cao S, Zhang Y, et al. Learning-based single-step quantitative susceptibility mapping reconstruction without brain extraction. NeuroImage. 2019; 202: 116064.

